# Prevalence of Late-Stage Parkinson’s Disease in the US Health Care System: Insights from TriNetX

**DOI:** 10.1101/2024.01.29.24301955

**Authors:** Sol De Jesus, Annika Daya, Liba Blumberger, Mechelle M. Lewis, Doug Leslie, Samer D. Tabbal, Rachel Dokholyan, Amanda M. Snyder, Richard B. Mailman, Xuemei Huang

**Affiliations:** Departments of Neurology, Pennsylvania State University-Milton S. Hershey Medical Center, Hershey, PA, USA; Public Health Science, Pennsylvania State University-Milton S. Hershey Medical Center, Hershey, PA, USA; Pharmacology, Pennsylvania State University-Milton S. Hershey Medical Center, Hershey, PA, USA; Radiology, Pennsylvania State University-Milton S. Hershey Medical Center, Hershey, PA, USA; Neurosurgery, Pennsylvania State University-Milton S. Hershey Medical Center, Hershey, PA, USA; Translational Brain Research Center, Pennsylvania State University-Milton S. Hershey Medical Center, Hershey, PA, USA; Department of Kinesiology, Penn State University, University Park, PA 16802 USA

**Author notes:** These authors contributed equally (Sol De Jesus, Annika Daya, Liba Blumberger). **<u>Corresponding Authors</u>** Sol De Jesus, MD, Department of Neurology, Penn State-Hershey Medical Center, Hershey, PD 17033-0850, Xuemei Huang, M.D., Ph.D., Department of Neurology, Penn State University, H037, 500 University Drive, Hershey, PA 17033-0850.

**Keywords:** Parkinson’s disease, late-stage Parkinson’s disease, health care burden, palliative care

## Abstract

**Background:** Patients in late-stage Parkinson’s disease (PD_LS_) are caregiver dependent, have low quality of life, and higher health care costs.

**Objective:** To estimate the prevalence of PD_LS_ patients in the current United States (US) health care system.

**Methods:** We downloaded the 2010-2022 data from the TriNetX Diamond claims network that consists of 92 USA health care sites. PD was identified using standard diagnosis codes, and PD_LS_ was identified by the usage of wheelchair dependence, personal care assistance and/or presence of diagnoses of dementia. Age of PD_LS_ identification, and survival information are obtained and stratified by demographic and the disability subgroups.

**Results:** We identified 1,031,377 PD patients in the TriNetX database. Of these, 18.8% fit our definition of PD_LS_ (n=194,297), and 10.2% met two or more late-stage criteria. Among all PD_LS_, the mean age of PD_LS_ identification was 78.1 (±7.7), and 49% were already reported as deceased. PD_LS_ patients were predominantly male (58.5%), with similar distribution across PD_LS_ subgroups. The majority did not have race (71%) or ethnicity (69%) information, but for the available information, >90% (n=53,162) were white, 8.2% (n=5,121) Hispanic/Latino, 7.8% (n=4,557) black, and <0.01% (n=408) Asian. Of the PD_LS_ cohort, 71.6% identified with dementia, 12.9% had personal care assistance, and 4.8% were wheelchair bound.

**Conclusions:** Late-stage patients are a significant part of PD landscape in the current US healthcare system, and largely missed by traditional motor-based disability staging. It is imperative to include this population as a clinical, social, and research priority.

## Introduction

Parkinson’s disease (PD) is characterized clinically by motor dysfunction that includes bradykinesia, tremor, and rigidity. The 20^th^ century witnessed tremendous advances in understanding the role of nigrostriatal system in PD,^1-4^ as well as developing therapeutic strategies targeting at normalizing it.^5-11^ The first breakthrough was the availability of levodopa that effectively restored the lost dopaminergic tone.^2^ This led to a new era for PD patients with dramatically improved symptoms and lengthened lifespan.^5-11^ As of the first quarter of 21st century, PD patients remained largely independent in the first decade of their disease, especially when treated in PD subspecialty clinics and research centers.^12^

PD patients suffer from increasing motor (e.g., motor fluctuations, levodopa-induced dyskinesia, gait disorders, and falls)^13^ and non-motor (e.g., cognitive, mood, and autonomic) signs and symptoms.^14,15^ Together, this impacts overall independence and quality of life (QoL).^14,15^ It is estimated that PD affects at least six-million people worldwide,^16^ a number that may double globally by 2040.^16^ A recent GBA report suggested that the increasing PD prevalence has outpaced all other neurological disorders, akin to a pandemic.^17^ The increase has been attributed to several factors including an aging population, improved symptom detection, decline in smoking prevalence,^18^ and a variety of environmental perturbations,^19-22^ although much remains to be learned.^17,18^

PD is diagnosed based on clinical signs and symptoms. The relentless progression of PD has been tracked with Hoehn and Yahr (H&Y) staging that focuses on limited motor and postural instability milestones.^5,23^ With the widespread use of levodopa (the standard of care for more than 50 years) a robust clinical drug response became one of the criteria for consideration of clinical diagnosis of PD, with definitive diagnosis relying on the pathological presence of α-synuclein (αSyn)-positive Lewy bodies/neurites and dopamine neuron loss in the substantia nigra pars compacta (SNc) of the BG.^3,4^ Despite recent awareness of extra-nigrostriatal involvement and non-motor disabilities, PD staging has still largely focused on HY staging. This was the motivation for a recent Movement Disorders Society-sponsored meeting that addressed the many issues involved in developing a more precise PD staging system.^24^

Coelho and Ferreira^15^ more recently proposed a very useful operational definition to depict PD clinical progression. They labeled early-stage PD (PD_ES_) as beginning with initial diagnosis that is marked by substantial motor improvement when patients take levodopa; this is often called the “honeymoon” period because most patients have excellent QoL. Patients then progress to advanced-stage PD (PD_AS_) that is marked by levodopa-related motor complications (e.g., motor fluctuations and drug-induced dyskinesia). For this stage of patients, deep-brain stimulation (DBS) attenuates symptoms and can improve QoL dramatically. Finally, they^15^ proposed an operational definition of late-stage PD based on a Schwab and England Scale.^25^ Patients enter late-stage PD (PD_LS_) marked by levodopa-unresponsive postural instability, dysphagia, prominent autonomic dysfunction and/or dementia, and the need for assistance in walking or even being bed-bound. This was related to Schwab and England^25^ scores of less than 50% during periods of adequate symptom control (“on” period), described a patient requiring help with half of their chores and experiencing difficulty with all activities, and a score of 40% reflected being highly dependent on caregiver support, able to assist with all chores, but being unable to complete most tasks alone.^15^ Others have felt this conceptualization was useful (e.g.,^14 26,27^).

The late stage of disease (i.e., PD_LS_) has received less attention in clinical research, partly due to the complexity of clinical symptoms and other obstacles such as limited ability to travel. Late-stage disability may emerge as early as 7-10 years after diagnosis,^14,15,28,29^ and leads to PD_LS_ patients having increased cost and greater utilization of healthcare systems.^30^ Analysis of Medicare claims-based data has provided a picture of the financial impact of advanced/late PD (incurring greater mean costs) as compared to mild or moderate PD patients.^31^ In addition to falls and dementia, PD_LS_ often is characterized by dependence on caregivers for activities of daily living. Wheelchair dependence and institutionalization are also common. These disability milestones have been suggested to precede death in PD patients by ∼5 years.^15,32^ The exact prevalence of patients in later stages is, however, unknown, thus hindering effective need assessments for clinical, research, and social planning.

Leveraging the available TriNetX Diamond Network data, we have estimated PD_LS_ prevalence in the U.S health care system. We identified all PD patients, and from these data, determined those in late stages based on the disability milestones of wheelchair dependence, dementia, and/or personal care assistance. We then identified the characteristics of the PD_LS_ patient population to analyze their profiles.

## Methods

We utilized the TriNetX Diamond Network, a de-identified clinical patient dataset from a network of healthcare organizations in the US.^33^ This database provides ambulatory and primary care electronic medical records, medical claims, and patient medication data from 92 healthcare organizations in the US and includes data for 212-million patients. According to the TriNetX website, the Diamond Network contains data covering 1.8 million providers and 99% of US health plans. The data covered a time window of January 1 2010 to the date of data extraction on February 24, 2023, and then identified and described individuals with PD.

A preliminary query was performed on the TriNetX website to look at the overall PD cohort using the ICD-10 and ICD-9 diagnostic codes entered in an electronic medical record (EMR) to specify PD in clinical practice (G20; 332.0). PD_LS_ was defined as requiring a diagnosis of PD plus one or more of the following mentioned disability milestones. The presence of PD_LS_ was identified via TriNetX query tool with the following combinations of ICD-10 and ICD-9 (Table 1) diagnostic codes: a diagnostic code for PD (G20; 332.0) in combination with a code for wheelchair dependence (Z99.3; V46.3), dementia (F02.80; F02.81; 294.11; 294.10), and/or personal care assistance (Z74.1; Z74.2; Z74.3; V60.89; V60.4). It is noted that the personal care assistance codes do not specify the degree of caregiver support but provides a good initial indicator of the need for personal care assistance. Because PD diagnosis before age 40 is rare (often with atypical features and progression), those individuals were excluded although this may have eliminated some patients with young onset PD. For transparency and better understanding of source data, we also captured all data, even if incomplete or with otherwise inaccurate data entry (Figure 1).

**Figure 1:**
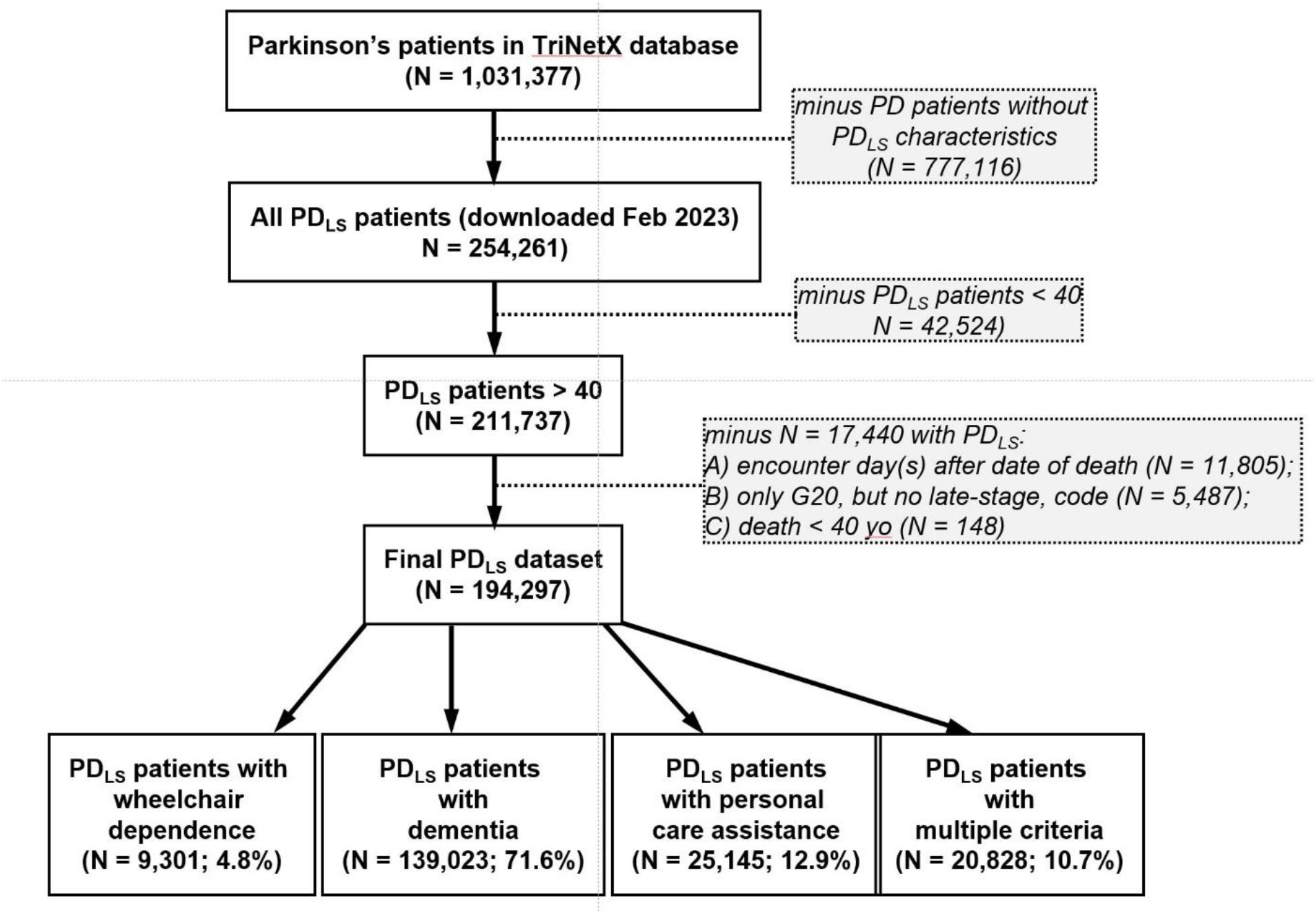
PRISMA Flow Diagram of Cohort Exclusions, Data from the TriNetX database. Data were downloaded and included information from patients with PD_LS_. Since this study focused on the late-stage characteristics of patients with Parkinson’s disease, we excluded patients without PD_LS_ characteristics, those aged <40, and those where the reported date of death occurred prior to a documented encounter. Patients then were separated by the PD_LS_ characteristics that included patients with wheelchair dependence, dementia, personal care assistance, or a combination of these (see Figure 2).

The resulting data library for PD_LS_ patients was requested for the following additional analyses. First, individual PD_LS_ subgroups, based on our querying criteria, were extracted without overlap (PD + one late-stage criterion). Second, we created combined PD subgroups to capture the presence of multiple late-stage criteria (multiple criteria groups) encompassing all possible combinations: (a) dementia and wheelchair; (b) wheelchair and personal care; (c) dementia and personal care; and (d) presence of all three criteria. These data are presented in a Venn Diagram (Figure 2). National Drug Codes (NDCs) were used to identify drug usage associated with Parkinson’s disease as levodopa-related, other dopaminergic, memory enhancing, and or antipsychotic medications (see Supplemental Data for details).

**Figure 2.**
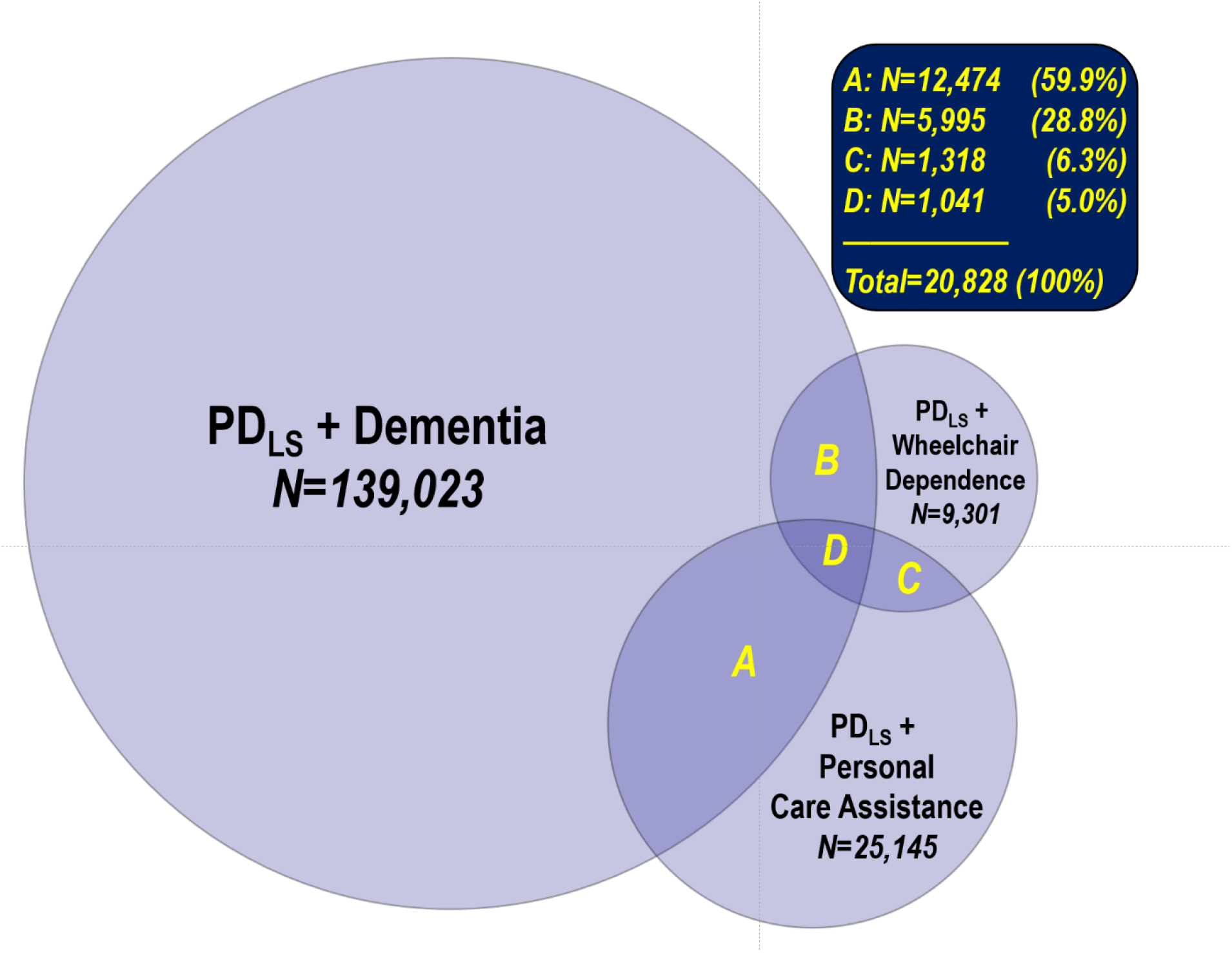
Venn diagram describing overlap in diagnoses and late-stage disease criteria among the PDLS patients. Patient numbers in each overlapped are inclusive only of those with the listed combination. That is, section A (12,474 patients in PD_LS_+Dementia vs. PDLS+Personal Care Assistance) does not include N= 1,041 who also have wheelchair dependence.

A non-parametric test, chi-square, was conducted to assess differences in observed frequencies across stratified groups. All analyses were conducted using SAS Version 9.4, and final data was drawn February 24, 2023. Penn State Institutional Review Board prior approval was unnecessary due to the de-identified nature of the database. Descriptive statistics were used to summarize sample characteristics with frequencies computed for demographic, prescription, and death data that were stratified by late-stage criteria (dementia, wheelchair dependence, personal care assistance, and their combinations). Binary variables were created to identify whether an individual was alive or deceased at the time of data retrieval. Age was calculated by subtracting a patient’s year of birth from their last service date documenting an interaction with the healthcare system partnering with TriNetX. All descriptive analyses were conducted using SAS Version 9.4, and final data was drawn February 24, 2023. Penn State Institutional Review Board prior approval was unnecessary due to the de-identified nature of the database.

## Results

### Identification of the PD_LS_ cohort and disability distribution in TriNetX dataset

From the preliminary TriNetX data query, 1,031,377 patients between 2010 and 2022 had a diagnosis of PD. Within the overall PD group, 777,116 subjects did not have any PD_LS_ features. After excluding 42,524 subjects who were < 40 yo, 211,737 PD_LS_ patients > 40 yo remained. We also excluded another 17,440 patients: these include patients with an encounter day(s) documented after the reported date of death (N = 11,805); those with only a G20, but no other late-stage code (N = 5,487); and those who died before age 40 (N = 148). This gave a final dataset of 194,297 PD_LS_ patients (Figure 1).

Of the total PD_LS_ cohort, more than 70% (139,023) were assessed as having dementia, 9,301 subjects were identified as having wheelchair dependence, 139,023 subjects were identified with a diagnosis of dementia, and 25,145 subjects were specified utilization of personal care assistance (Figure 2). Among PD_LS_ patients, 10.2% (n=19,787) had at least two of these other late-stage criteria (Figure 2). Within those with multiple diagnoses, 3.1% (n=5,995) had both the diagnosis of dementia and wheelchair dependence; 0.7% (n=1,318) were wheelchair dependence and required personal care assistance; 6.4% (n=12,474) had dementia and personal care assistance, and 0.5% (n=1,041) met all three criteria (Figure 2).

### Age and vital status of PD_LS_ cohort and its disability subgroups in TriNetX dataset

The mean age was 77.9 ± 7.7 at the time of PD_LS_ identification in TriNetX database (Table 2). The mean age was slightly lower for those having wheelchair dependence (74.7 ± 9.5 y) and personal care assistance (74.7 ± 7.1 y). The mean age was higher in those diagnosed with dementia (78.9 ± 7.1 y) and those with multiple diagnoses (78.1 ± 7.6 y).

By February 2023, 45% percent (n=88,071) of all PD_LS_ were reported to be deceased. About half of PD_LS_ patients with dementia had died at the time of analysis (49.2%, n=68,368) followed closely by those with multiple disabilities needs/diagnoses (43%, n=9,027). Those with only wheelchair dependence (36.2%, n=3,369) or personal care assistance (29.1%, n=7,307) had fewer deaths (Table 2). For those who had died, the average age at death was 81.8 ± 6.6 y, similar across all groups although slightly lower in the Wheelchair and Personal Assistance groups (Table 2)

### Demographic characteristics of PD_LS_ cohort and disability subgroups in TriNetX dataset

A majority of the PD_LS_ patients were male [58.4% (n=113,406) versus 41.6% female (n=80,812) (Table 2)]. The pattern of male predominance seemed to persist in all PD_LS_ subgroups, except for those with PD_LS_ + wheelchair dependence that were sex equivalent. Race was unknown for 71.9% (n=139,630) of PD_LS_ patients; 25.8% (n=50,035) were documented as White, 2.2% (n=4,236) as Black or African American, and 0.2% (n=396) were Asian. Similarly, ethnicity was reported sparsely for PD_LS_ patients: there were no reports for 69.9% (n=135,812); 27.6% (n=53,635) reported as not Hispanic or Latino, and only 2.5% (n=4,850) Hispanic or Latino. [Statistical analyses are in Supplemental tables]

### Use of antiparkinson and adjuvant drugs in PD_LS_

We queried the database for use of antiparkinson and related adjuvant drugs in PD_LS_ (see Methods and Supplemental Information for details). Slightly more than 60% of the people we identified as PD_LS_ were using <u>none</u> of the drugs we have categorized as providing symptomatic relief to Parkinson’s patients; these drugs include levodopa in any formulation, dopamine agonists, or MAO inhibitors or cholinesterase inhibitors (Table 2).

## Discussion

Lewis et al.^12^ recently highlighted the current landscape of PD progression in the early 21^st^-century, especially noting that PD patients remain largely independent in the first decade of disease. There is, however, a paucity of data and clinical metrics capturing the real-world PD progression in patients beyond 10 years, and/or in later stages of disease. Here, we have used the TriNetX clinical/claims database to capture and characterize patients in late-stages of disease (PD_LS_), using an operational definition advocated by Coelho and Ferreira.^15^ Our findings suggest a significant proportion of PD patients (∼20%) meet criteria for PD_LS_. This is consistent with more limited studies both within^31,34^ and outside the US.^30^Given that the number of PD patients is expected to double globally by 2040, there likely will be dramatic increases in both the prevalence and healthcare cost that occur for people with PD_LS_.^30^ Thus, it is imperative to include late stage PD population as clinical, social and research priority as a potential and emerging health emergency.

### PD_LS_ as a new clinical stage entity: guide the assessment of clinical and society needs

With improved pharmacologic treatment, advanced therapies, and increasing access to care, patients with PD are living longer and reaching later stages of the disease. In this study, we estimated the prevalence of the PD population in the late-stages, guided by an operational definition advocated by Coelho and Ferreira.^15^.

Our current data underscores the limitation of a motor-centric staging, as embodied by H&Y staging, by demonstrating its inadequacy to capture greater than 95% in late stages with unmet needs. Our data also pointed out that dementia is composed of the largest group (>75%) of PD patients in the late stage, this number is consistent with the dementia prevalence observed by other groups.^14,35-37^ Recent studies clearly suggested that those with PD-normal cognitive function may have a normal life expectancy, whereas those with PD-MCI or PD-dementia may have reduced life expectancy.^38^ Together, our findings support the clinical and research priorities of dementia in PD, converging with the current NIH priority of Alzheimer’s disease and related disorders.

As the research on better staging terminology is still underway for PD patient,^24^ clinical coding practices vary significantly among providers. There is a lack of standardization as well as non-specific coding for PD to capture disease-associated manifestations. Dashtipour et al.^39^ have highlighted the limitations that exist with the current ICD-10-CM coding for PD, particularly in delineating motor complications with disease progression. Inclusion of disability milestones separately from the standard PD diagnostic code (G20) can be an initial way to capture more accurately the patient status and disease severity. With improved capability of tracking larger populations afforded by the expansion and accessibility of claims-based databases (e.g., TriNetX), accurate coding becomes integral to understand the impact of interventions and management and informing research on PD subpopulations like PD_LS_.

According to a recent global health (GBA) report, PD is the fast growth neurological disorder, outpaced all other neurological disorders, including AD and related disorders (ADRD).^40^ PD_LS_ patients are anticipated to have increased both prevalence and health care system utilization, highlighting the urgent need for improved identification, and tracking of this proportion of PD patients in the current health care system. The results will help guide more equitable, and impactful health care resource allocation.

### PD_LS_ as a biomedical opportunity: an understudied research population

Current research efforts focus primarily on early-stage PD. The new data shed light on the importance and urgency of integrating efforts to reduce the late-stage disease burden into current research paradigms. As we have noted,^12^ the treatment options for symptom relief, disease-modifying therapies, and neuroprotective interventions have continued to evolve.^41^ Most of the existing approaches, however, are not focused on PD_LS_ patients with the exception of parenteral administration of levodopa/carbidopa formulations. The latter has shown consistent but modest effectiveness and can be associated with significant adverse events based on the invasive nature of the procedure.^42-44^ Moreover, PD_LS_ patients are likely to have had dopamine terminal degeneration such that processing levodopa adequately is no longer possible.^13,45-48^ Thus, PD_LS_ patients may forego specialized care under the assumption there are limited care options or available pharmacologic treatments.^14^

Recent publications from others raise awareness that late-stage PD patients is an important and neglected group.^49-55^ The care for PD_LS_ patients shifts to providing support and comfort, often with poor QoL. This underscores the need for multidisciplinary individualized treatment goals and identifying appropriate pharmacologic and nonpharmacologic interventions carefully chosen from existing toolsets for late stage PD patients.^56^ Similarly, PD_LS_ patients are usually excluded from pharmaceutical trials. Of the available treatments, most are pharmacological and targeted at dementia (e.g., rivastigmine, donepezil, galantamine) or dopaminergically-induced psychosis (e.g., clozapine, pimavanserin) with modest levels of success.^20,57,58^

Current research has yet to address the needs of PD_LS_ patients. Deep brain stimulation is an established surgical treatment option for patients with troublesome motor fluctuations, responsive to levodopa and without dementia.^59,60^ PD_LS_ patients who have undergone DBS also experience disability PD_LS_ milestones, and are excluded from, or considered ineligible, for PD research.^61,62^ In clinical practice, many PD_AS_ and PD_LS_ patients with/without DBS and their caregivers still look for treatment and research opportunities.^13,63^ Although, potential disease-modifying therapies and neuroprotective interventions in early and even earlier (prodromal stages)^64,65^ continue to be sought..

Finally, we noted that >60% of the PD_LS_ population were using none of the drugs common for treating PD including levodopa. This finding is both novel and surprising because levodopa is gold-standard for treating PD, even only for symptomatic relief and incrementally for comfort care. The exact cause for this finding is unclear, one hypothesis is that the near total loss of striatal loss of dopamine innervation in PD_LS_ causes levodopa conversion to dopamine in the striatum to be insufficient, rendering levodopa ineffective and/or causing intolerable side effects due to off-target effects.^13,66,67^ Similarly, available D_2/3_ dopamine agonists have only modest antiparkinson effects and dose-escalation is not possible because of extrastriatal side effects. It also is possible that a subset of these patients may have been misdiagnosed, and have a parkinsonism like MSA, PSP, of LBD rather than PD.^68^ Because of the more rapid clinical progression and the lower prevalence of atypical parkinsonism, it is likely they constitute only a small minority of the 60%, but detailed examination would be worthwhile. Another possibility is that some portion of the 60% of patients not using antiparkinson drugs was the result of missing data as often occurs in large databases.

### Limitations and future directions (

We are cognizant of several limitations of our analyses. Inherent to real world claims-based datasets, significant demographic information (e.g., race, ethnicity) were under-reported. Indeed, there was no race and ethnicity information for two-thirds of the subjects, making it impossible to determine if a specific underserved population could be identified.^69^ Given no current formal definition for PD_LS_ the operational definition utilized is restricted to three specific disability milestones. Moreover, there are inaccuracies in data entry and limitations in the diagnostic codes for PD and PD-associated symptoms within EMR systems.^11,70,71^ This is emphasized in utilizing codes such as personal care assistance where degree of assistance necessary is not detailed. Future coding iterations may benefit by providing more granular details to reflect patient status. We also could not estimate the exact data of PD symptoms onset or diagnoses as the subtle motor- and/or non-motor symptoms may occur earlier than the date when patients sought professional advice that would have been into the TriNetX. There also is a lack of data on the subspecialties (a movement disorder specialist), if any, that made the PD diagnoses. Additionally, certain diagnostic codes may have been under-utilized, contributing to under-estimates of disease progression, although the driving factors for such practices are unclear and should be studied further to improve EMR data capture.

Other cognitive disability milestones such as psychosis/hallucinations were not included in this analysis because heterogeneity and limitations inherent in the definition and coding practices likely are amplified in such databases.^72^ These data were obtained from multiple health care organizations in the US, but may not fully represent the overall PD population since a third of the US population was not captured by this EMR database. Our data also suggest that there may be either unequal access to healthcare, or markedly under-reported prevalence of PD, in several racial or ethnic groups, an issue that requires attention. Therefore, the number of PD_LS_ estimated based on ICD codes in our study is certainly an underestimate.

## Conclusions

The prevalence of PD_LS_ is significant, and thus its impact will increase markedly in the future. Our data indicate that the levodopa-based treatment regimens may lose utility in PD_LS_, as more than 60% of these patients are no longer taking antiparkinson drugs. Together, these data are more evidence of this large and underserved population in our system. Future clinical and research priorities should include the PD_LS_ to improve understanding of healthcare utilization patterns, medication status, cost-burden, and intervention-based outcomes. These insights will be meaningful in clinical setting as we can choose the current most effective therapies, either pharmacological, surgical, and supportive, for PD_ES_, PD_AS_ and PD_LS_, respectively. Scientifically, we should embrace the challenges of a growing presence of PD_LS_, capture the potential of emerging novel dopaminergic agents,^13,63,66,73^ and devise future strategies to address these unmet needs. The implementation of new diagnostic codes for late-stage patients may provide simpler and defined disability milestones that encourage tracking of disease progression, inform health economics burden, and identify supportive treatment options in real world health care system.

## Supporting information

Supplement 2: Drug Codes searched

## Data Availability

All data produced in the present work are contained in the manuscript

## Disclaimer

Dr. Lewis now is employed by the National Institute of General Medical Sciences (NIGMS). Her role in this study was as an employee of Penn State University. The opinions expressed in this article are those of the authors, and do not reflect the views of the National Institutes of Health or any other external organization.

